# Perceptions of parenting twins as experienced by fathers: A qualitative study

**DOI:** 10.1101/2025.10.05.25337379

**Authors:** Maki Kanzaki, Hiroko Sakai

## Abstract

This study aims to qualitatively explore how fathers raising twins acquire their paternal roles and how their perceptions and attitudes change throughout the parenting process. The study was conducted through online semi-structured interviews with 10 fathers of twins, based on an interview guide. A qualitative inductive method was employed to analyze and structure fathers’ perceptions of childcare. An exploratory net was created from 388 labels, following which categories were derived through multi-stage pick-up, and groups were formed and illustrated. The resulting structure illustrated a dynamic process in which fathers, who were unable to fully comprehend what it meant to become a father of twins during pregnancy, confronted the unexpected realities of twin parenting after birth. This led to not only psychological and emotional challenges, but also gradual adaptations and transformations. The findings underscore the need for early interventions to support fathers and contribute to the development of comprehensive support systems that consider the complexities of parenting twins. In the future, developing phased support programs for fathers— beginning in the prenatal period—and promoting broader social understanding involving families, communities, and workplaces will be essential in fostering environments that enable sustainable work–life balance for fathers.

## Introduction

Japan’s declining birthrate continues to present a major demographic challenge, with the total fertility rate reaching a record low of 1.20 in 2023 [1]. Moreover, the average age of childbearing has increased, as has the use of assisted reproductive technology (ART [2]. Following the inclusion of ART in the national health insurance system in 2022 [3], its utilization has grown rapidly, with the incidence of twin births expected to rise further [1]. This demographic shift raises concerns regarding perinatal complications associated with multiple pregnancies and the substantial parenting burden they entail [4]. Notably, fathers of twins are believed to experience particularly high levels of psychological stress [5,6].

Approximately 10% of fathers in Japan have been reported to exhibit postpartum depressive symptoms [7]. Identified contributing factors include the burden of parental roles, mental health status, and the quality of marital relationships [8,9], with these risks potentially being exacerbated in the context of parenting twins [5,6]. However, research focusing specifically on fathers of twins is scarce. Although a few studies have examined their parenting experiences and mental health [5,6,10–12], little is known about their specific support needs or the unique challenges they face. Most prior studies have examined fathers of singletons or mothers of twins, resulting in a lack of in-depth understanding from the paternal perspective.

Parenting twins involves distinct challenges not encountered in singleton parenting. These challenges may influence the development of paternal identity and the process through which fathers internalize and enact their parenting roles [10,13]. Understanding how fathers of twins perceive, negotiate, and evolve in their paternal roles—particularly in the face of unique emotional and practical difficulties—can provide valuable insights to enhance family support systems and formulate effective parenting policies.

Therefore, this study aims to qualitatively explore how fathers raising twins acquire their paternal roles and how their perceptions and attitudes change throughout the parenting process. The findings are expected to not only establish foundational knowledge to guide the development of psychological and social support strategies for fathers of twins but also inform clinical and community-based interventions that may have relevance in international contexts.

## Methods

### Definition

In this study, “multiple pregnancy” is defined as a pregnancy in which two or more fetuses coexist in the uterus simultaneously[14]. A “multiple-birth family” refers to a family raising more than one child of the same age, such as twins or triplets [15]. The term “paternal role” is defined as having a sense of identity and responsibility as a father from the prenatal stage of a twin pregnancy; providing financial support; assisting the wife in adapting to her maternal role; cooperating in household and childcare tasks; discussing parenting policies; and adapting to the role of a father.

### Study design

This study employed a qualitative inductive research design.

### Theoretical framework

This study was conducted in parallel with research focused on the process of paternal role acquisition [16], and is theoretically grounded in Thornton and Nardi’s (1975) role acquisition model [17], according to which, individuals adapt to new roles through a process of both passive and active experiences. The acquisition of a role is characterized as a temporal process encompassing four stages—anticipatory, formal, informal, and personal—through which individuals move from passively accepting a role to actively internalizing and performing it [17].

In this study, the model was adopted to frame the experiences of fathers of twins from their wives’ pregnancies onward to understand how they acquired their paternal roles through the four stages of role acquisition.

### Participants

Eligibility criteria for participants included: being a biological father of twins, being 18 years or older, cohabiting with the wife, raising twins aged between 3 months and approximately 1 year at the time of the interview, and having a wife without severe perinatal complications. Fathers were excluded if they or their twins had serious illnesses or disabilities. Twin pregnancies are known to be high-risk for both mother and infants, and neonatal hospitalization is common [4]. To ensure participants had begun actively parenting and could reflect on their experiences meaningfully, only fathers of twins aged at least 3 months were included.

Additionally, limiting the age of twins to approximately 1 year allowed the study to focus on a consistent developmental stage. Previous studies have included fathers of older twins (e.g., 2–9 years old) [11] or of infants aged 4–5 months [12], which introduces variability. Furthermore, because autobiographical memory—the recall of past experiences—is shaped over time by personal values and life perspectives [18,19], this study aimed to capture subjective experiences, emotions, and reflections from the prenatal period through the first year postpartum, while maintaining temporal proximity for accuracy of recall.

### Sample size

In line with the study’s exploratory objective of structuring paternal perceptions of raising twins, a total of 10 fathers who met the inclusion criteria and provided informed consent were recruited as participants.

### Data collection

#### Recruitment

Participants were recruited using snowball sampling and through the cooperation of seven community organizations in Japan that support families with twins. Although 11 fathers initially registered, one was excluded because his twins were already in early childhood. Ultimately, 10 fathers of twins aged between 3 and 15 months participated. They resided in Kanagawa, Ibaraki, and Chiba prefectures in the Greater Tokyo Area, and in Osaka and Hyogo prefectures, approximately 500 km west of the capital.

#### Participants characteristics

Of the 10 participants, 3 had older children, while 7 were first-time fathers.

#### Interview content and literature review

To explore a wide range of paternal experiences, this study utilized a semi-structured interview approach. An interview guide was developed based on a comprehensive literature review and structured chronologically in alignment with the role acquisition model, focusing on the father’s developmental process from the time of pregnancy confirmation onward (Table 1).

**Table 1.**
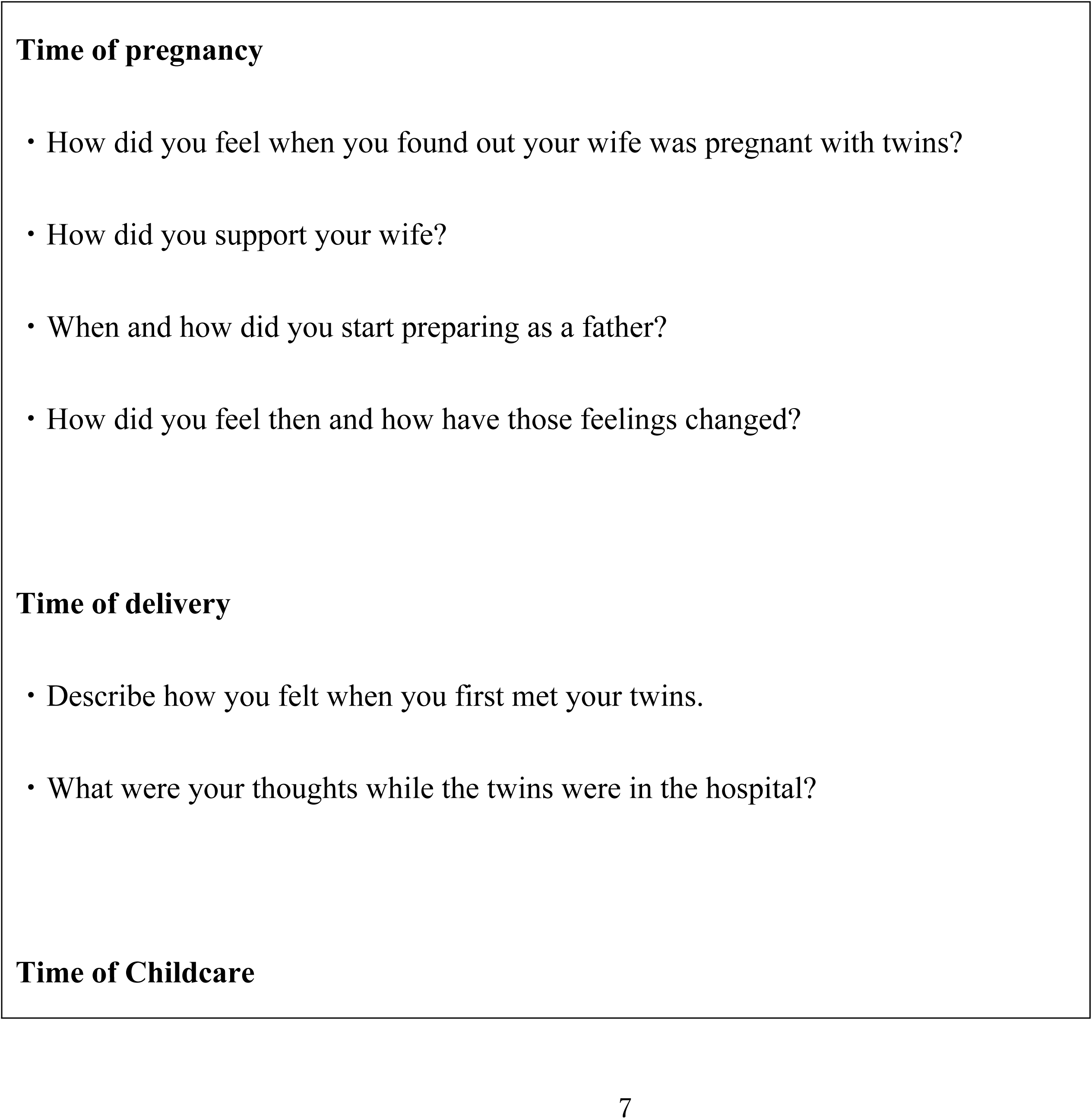
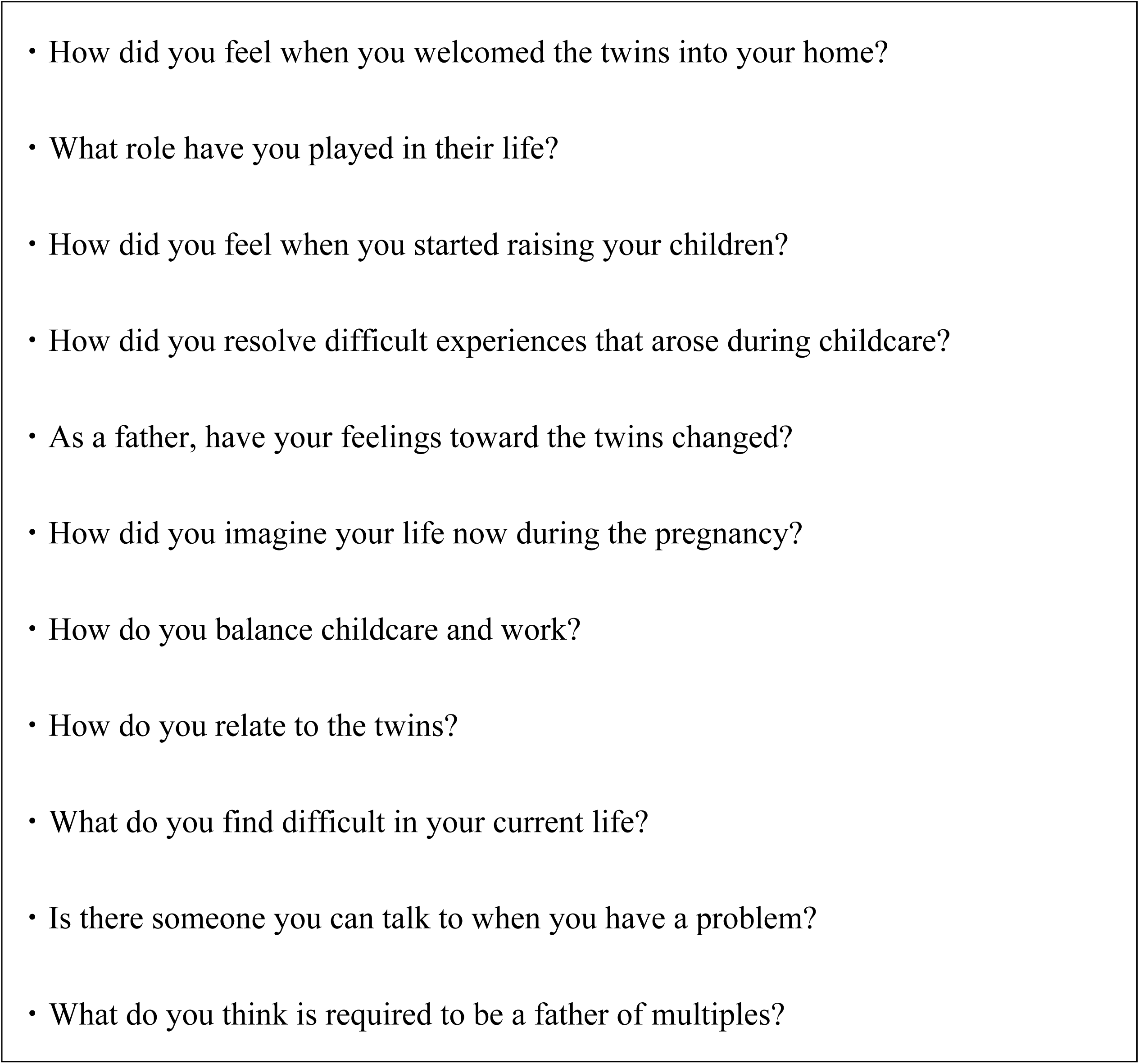
Interview Guide.

The literature review was conducted using PubMed, CINAHL Complete, ProQuest, and the Ichushi Web Ver. 5 databases. It sought to extract accessible peer-reviewed articles in Japanese and English published up to May 31, 2022. Commentaries, conference proceedings, and magazine articles were excluded. The review was guided by two key perspectives: (1) the developmental process of becoming a father and forming a parental identity, and (2) the unique experiences of fathers raising multiples (Figs 1 and 2).

**Fig 1.**
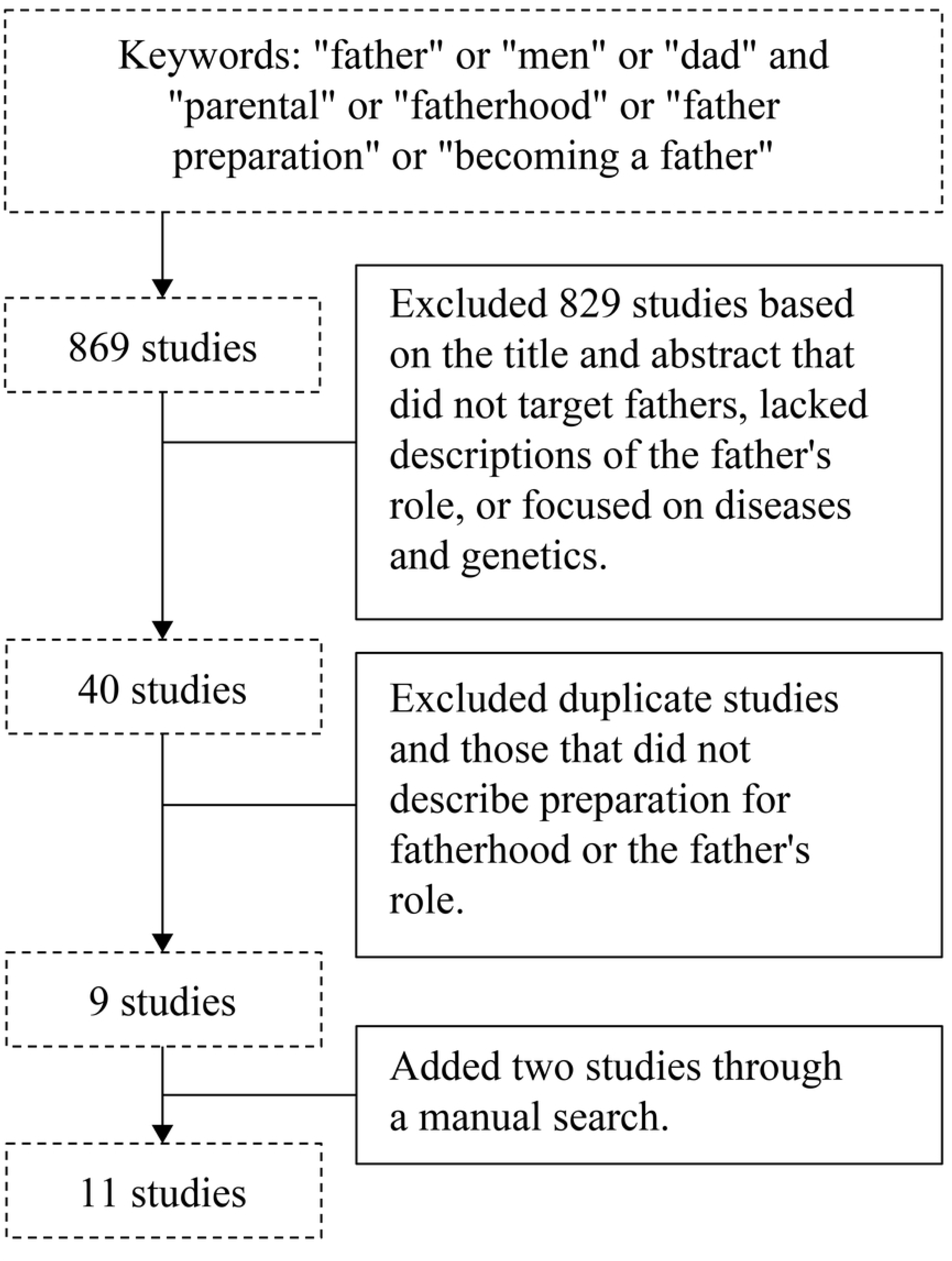
Literature Selection: Fatherhood Preparation and Parenthood.

**Fig 2.**
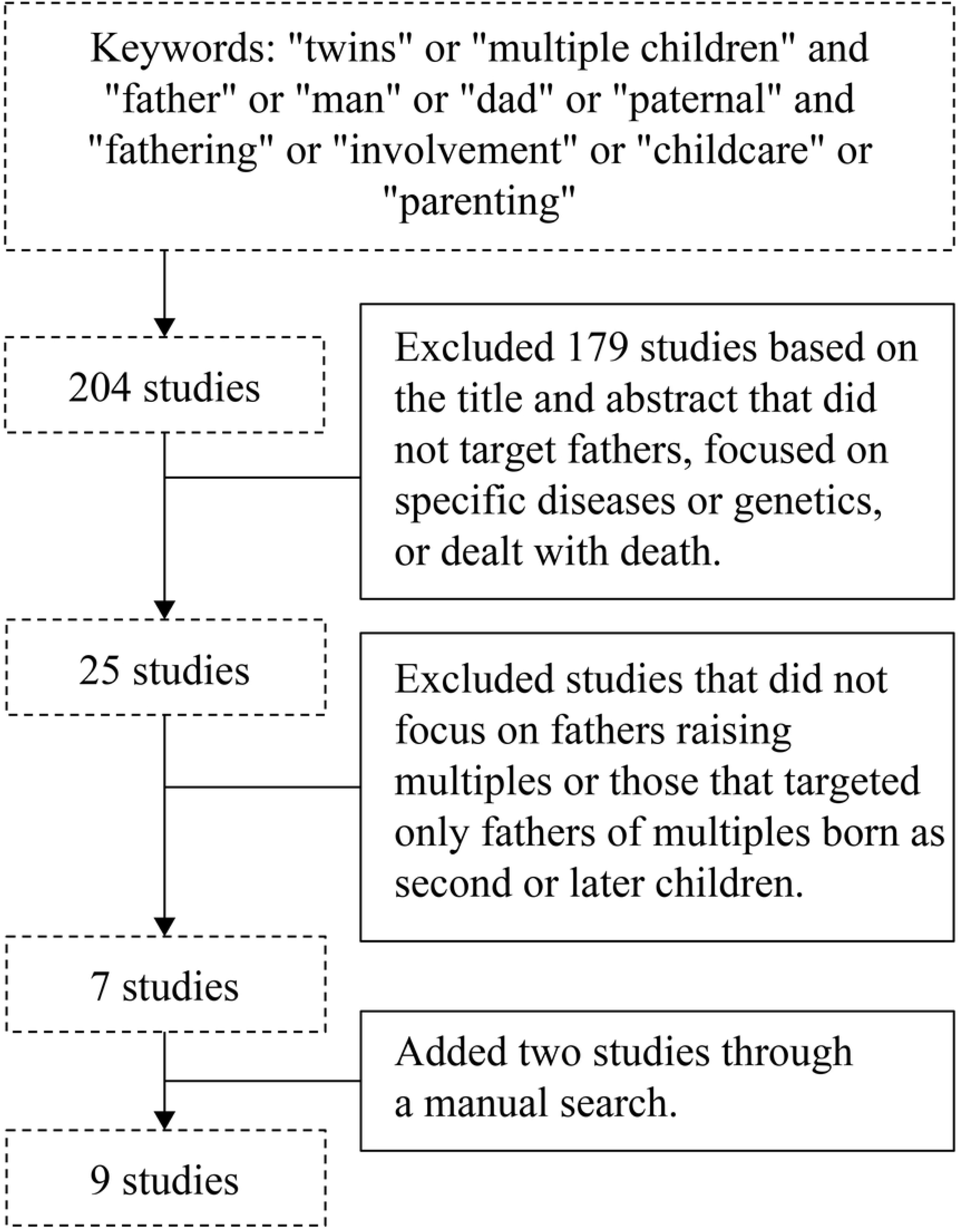
Literature Selection: Fathers Raising Multiples.

Becoming a parent is a process that involves the evolution of one’s personality through engagement in childcare roles [20,21]. For a man, this process begins with his partner’s pregnancy confirmation, which often triggers the development of paternal awareness [22–24]. Positive emotions during pregnancy and childbirth are associated with an increased sense of becoming a father [22,23], although men may also experience confusion, anxiety, or helplessness in response to their partner’s physical and emotional changes [20,22–25]. Nonetheless, feelings of affection toward their partner, joy in caregiving, and the gradual realization of their paternal identity help them adapt to the fatherhood role [22–26]. Moreover, cooperative parenting with the mother has been shown to promote paternal development [20,25,27].

Compared to fathers of singletons, fathers of multiples are more likely to experience childcare-related stress, anxiety, and depressive symptoms—owing to not only increased caregiving demands but also the use of ART [4,5,28,29]. Sleep deprivation and fatigue are particularly pronounced among fathers of twins [6,30], highlighting the importance of family and social support. In Japan, the use of paternity leave remains low because of concerns about its impact on employment status and career advancement [31,32].

This review contends that despite fathers of twins undergoing similar developmental processes in preparing for parenthood as those of singletons, the unique burdens of raising twins can pose significant physical and emotional stress. Therefore, understanding how these fathers perceive and engage in childcare may provide valuable insights that can inform the development of tailored support systems.

The interview guide was reviewed among the research team and validated by a qualitative research expert. A pilot test was conducted with two fathers of singletons (with 3- and 6-month-old infants, respectively) to assess the clarity and effectiveness of the items. Based on their feedback, unclear or ineffective items were removed. Data from the pilot test were not included in the final analysis.

#### Interview procedure

In light of the COVID-19 pandemic, all interviews were conducted online. Each interview was scheduled through email communication between the researchers and participants. With the participants’ consent, all interviews were audio-recorded using an integrated circuit recorder and later transcribed verbatim.

#### Study period

Participants were recruited between May 21 and August 23, 2022. Data collection was conducted from July through September 2022.

### Ethical considerations

Prior to participation, all participants received detailed written and oral explanations regarding the purpose of the study, the interview procedures, their right to voluntary participation and withdrawal, the protection of personal information and anonymity, and their right to access the study results. After providing written informed consent, they were given an additional verbal explanation immediately before the interview and confirmed their consent verbally at that time. This study was approved by the Medical Ethics Review Committee of Kansai Medical University (Approval No. 2021364).

### Data analysis

This study aimed to clarify two aspects: (1) fathers’ perceptions of raising twins, and (2) their views on what is expected of a father of twins. To structurally analyze these perceptions, we adopted the KJ Method, a qualitative analytical technique developed by Kawakita [33]. Although other qualitative methods—such as modified grounded theory and content analysis—are commonly used for systematic and inductive exploration (Saiki, 2013) [34], the KJ Method is particularly suited for this study because it allows researchers to extract structure and meaning from complex and often chaotic qualitative data by synthesizing patterns and themes. It has been widely applied across disciplines for its ability to provide a holistic understanding of a phenomenon [35,36].

The KJ Method was also selected to avoid imposing preexisting conceptual frameworks or researcher biases, instead allowing the voices of the participants to emerge directly from the data. Our goal was not to analyze individual cases in isolation, but to derive broader and transferable insights based on shared experiences.

The researchers had prior clinical experience as midwives in perinatal care for high-risk pregnancies and were familiar with family-centered care during pregnancy, childbirth, postpartum, and infancy. The primary analyst had previously received formal training in the KJ Method through certified workshops, including the basic, interviewing, and applied techniques courses, and continued attending follow-up workshops to deepen their expertise.

### Analytical procedures

The following steps were undertaken to analyze the data, as illustrated in Fig 3.

**Fig 3.**
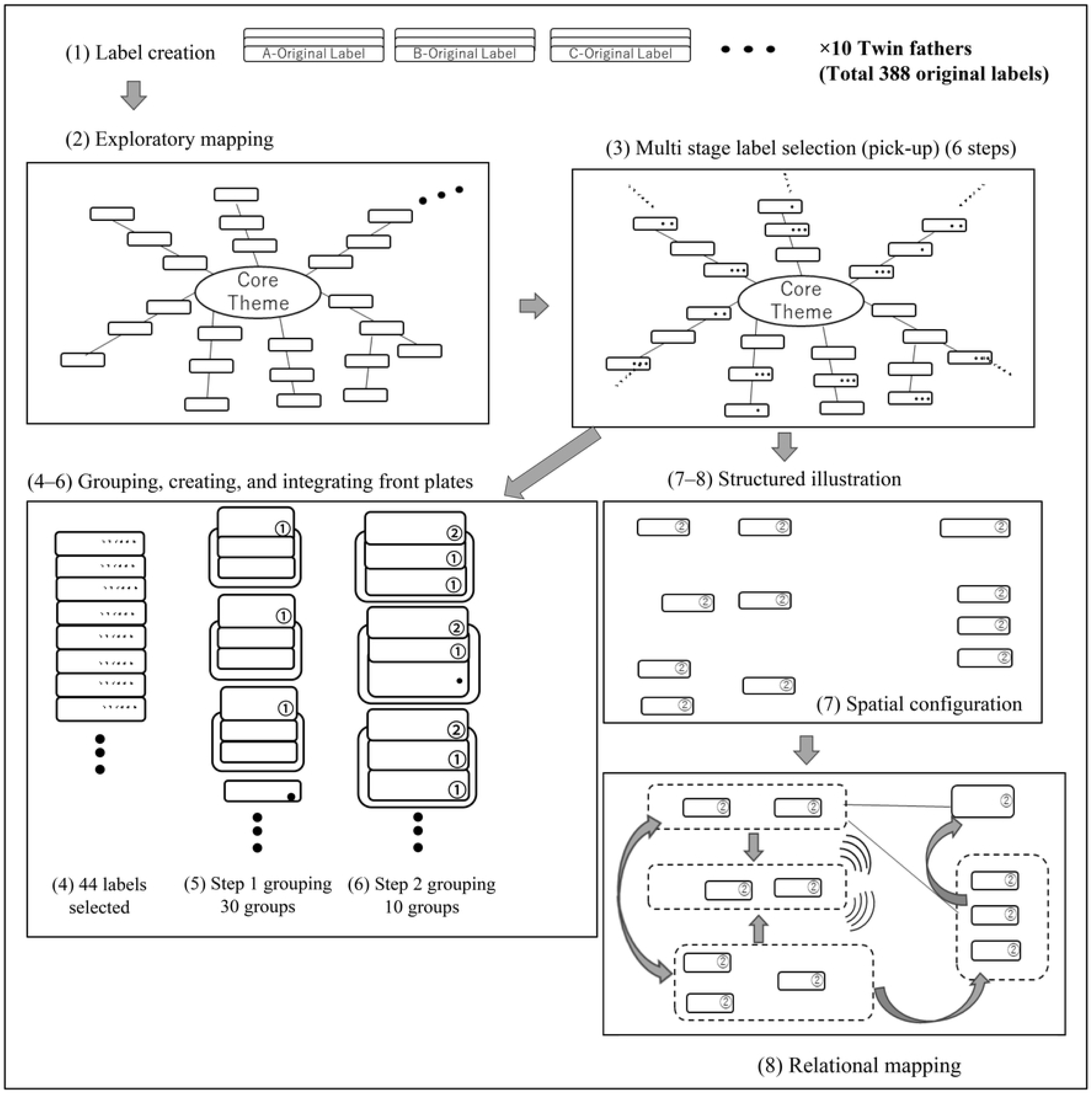
Flow of Analysis Following the KJ Method.

1. Label creation: In this step, transcripts were carefully read, and participants’ experiences related to raising twins—from the confirmation of pregnancy to the time of interview—were segmented and compressed into descriptive labels while preserving the original meaning.
2. Exploratory mapping: To assess the diversity of the data, a large sheet of paper was used to visually map labels radiating from the central theme, “fathering twins.” Related labels were grouped to identify overlapping, distinctive content.
3. Multi-stage label selection (pick-up): Labels were filtered based on their relevance and resonance (“What stands out?”) and gradually refined through multiple stages.
4. Group formation: Selected labels were expanded and grouped by similarity. Similarities within each group were verified and refined.
5. Title (Hyosatsu) creation: A core concept was abstracted from each group of labels and assigned a name plate as a title.
6. Integration through grouping: The titles were repeatedly synthesized into broader categories until approximately 10 titles remained, each representing a distinct thematic construct.
7. Spatial configuration: The final titles were arranged spatially to explore relational structures among them.
8. Relational mapping: Each cluster (“island”) was assigned a symbolic concept, and the relationships among islands were examined to develop a unified visual representation (Fig 4).

**Fig 4.**
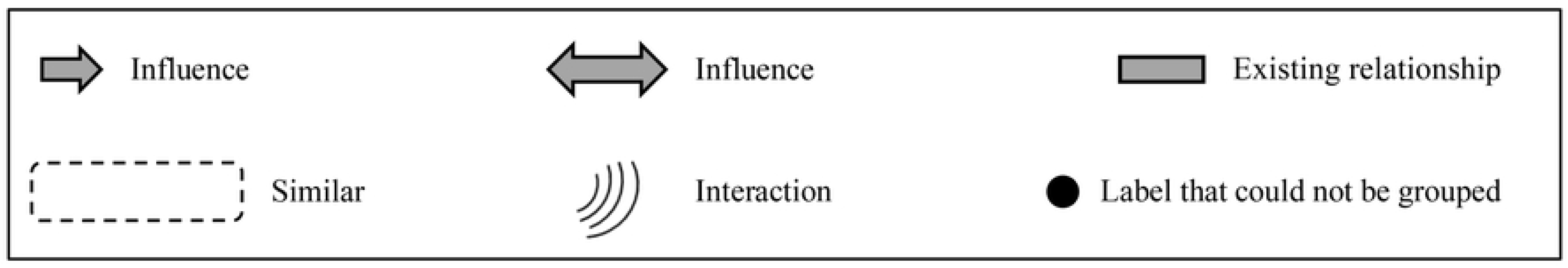
The KJ Method Relationship Line.

### Ensuring trustworthiness

Immediately after each interview, a transcript was created to understand what was conveyed and to confirm that no major revisions to the interview guide or sample size were needed.

Throughout the analysis process—from label creation to visual mapping—researchers held repeated discussions to share interpretations and reach consensus. The analytic process involved continually referring to the original transcripts to verify and revise labels and emerging structures.

To minimize interpretive bias, the research team received methodological guidance from Ms. Akiko Kawakita, a certified educator of the KJ Method. Furthermore, responses were rechecked with participants to ensure that the interpretations accurately reflected their experiences, thereby enhancing the credibility of the findings.

## Results

### Participant characteristics

The demographic characteristics of the 10 fathers who participated in this study are presented in Table 2. Interview durations ranged from 59 to 114 minutes. Seven participants were first-time fathers, with ages ranging from 26 to 42 years, and all fathers were employed. The types and durations of postnatal leave varied: five fathers took childcare leave, three took special leave, two used annual leave, and one used both special and annual leave. One father who did not take any official leave made personal adjustments to ensure time for parenting. Although six fathers reported that their partners were hospitalized owing to pregnancy-related complications, none experienced severe postpartum complications. Four fathers mentioned that their partners followed “satogaeri” (a customary return to the wife’s parental home during the perinatal period), and in two of these cases, the separation period between the fathers and their children lasted for two and ten months, respectively. Eight fathers received ongoing support from local governments, support organizations for families with multiples, or relatives.

**Table 2.** Summary of Participants.

|  | **A** | **B** | **C** | **D** | **E** | **F** | **G** | **H** | **I** | **J** |
| --- | --- | --- | --- | --- | --- | --- | --- | --- | --- | --- |
| **Age group** | 30s | 30s | 40s | 30s | 20s | 30s | 30s | 30s | 30s | 40s |
| **Occupation** | Office<br>worker | Office<br>worker | Office<br>worker | Medical<br>profession | Medical<br>profession | Medical<br>profession | Office<br>worker | Office<br>worker | Civil<br>servant | Civil<br>servant |
| **Years of**<br>**marriage** | 4 | 3 | 10 | 3 | 1 | 8 | 11 | 6 | 2 | 4 |
| **Wife's age**<br>**group** | 30s | 30s | 40s | 30s | 20s | 30s | 30s | 30s | 30s | 30s |
| **Wife's**<br>**employment** | Childcare<br>leave | Childcare<br>leave | Reinstated | Childcare<br>leave | Unemployed | Reinstated | Childcare<br>leave | Childcare<br>leave | Childcare<br>leave | Childcare<br>leave |
| **Wife's history** | Primiparity | Primiparity | Multiparity | Primiparity | Primiparity | Multiparity | Multiparity | Primiparity | Primiparity | Primiparity |
| **of childbirth** |  |  |  |  |  |  |  |  |  |  |
| **Number of**<br>**older children** |  |  | 4 |  |  | 1 | 2 |  |  |  |
| **Weeks of**<br>**conception**<br>**(weeks/day)** | 34/6 | 37/3 | 35/0 | 36/1 | 36/1 | 37/6 | 37/2 | 35/0 | 35/1 | 38/1 |
| **Birth weight**<br>**of first child**<br>**(g)** | 2048 | 2480 | 2158 | 1948 | 2200 | 3200 | 2532 | 2136 | 2360 | 2896 |
| **Birth weight**<br>**of second**<br>**child (g)** | 1962 | 2530 | 2438 | 2128 | 2230 | 2634 | 2646 | 1512 | 2450 | 2356 |
| **Number of** | 23 | 14 | 21 | 25 | 14 | 10 | 5 | 14 | 5 | 7 |
| **days of**<br>**child(ren)'s**<br>**hospitalization** |  |  |  |  |  |  |  |  |  |  |
| **NICU**<br>**admission**<br>**(both children**<br>**or one child)** | Yes | Yes | Yes | Yes |  |  |  | Yes |  |  |
| **Father's**<br>**participation**<br>**in prenatal**<br>**education** |  | Yes |  |  | Yes |  |  | Yes | Yes | Yes |
| **Duration of**<br>**leave for** | Time of<br>need | 1 year | Date of<br>birth and | Childcare<br>leave for 2 | Childcare<br>leave for 3 | 2 weeks of<br>special | 7 days<br>special | 41 days<br>childcare | 7 days<br>special | 3 weeks<br>childcare |
| **childcare and**<br>**type of leave** | (ineligible) |  | date of<br>child's<br>discharge<br>Annual<br>leave 2<br>Days | months<br>from 10<br>months of<br>age | months from<br>3 months of<br>age | leave and 1<br>month of<br>annual<br>leave | leave | leave | leave | leave |
| **Wife's**<br>**hospitalization**<br>**during**<br>**pregnancy** |  | Yes | Yes | Yes | Yes | Yes |  | Yes |  |  |
| **Method of**<br>**delivery** | CS | CS | CS | CS | CS | CS | V | CS | V | CS |
| **Gender** | F/M | M/M | M/F | F/M | M/M | F/F | F/M | M/F | F/F | F/F |
| **combination**<br>**of twins** |  |  |  |  |  |  |  |  |  |  |
| **Whether or**<br>**not the twins**<br>**were born at**<br>**home** |  |  |  | Yes | Yes | Yes |  |  |  | Yes |
| **Paternal and**<br>**paternal**<br>**separation**<br>**owing to**<br>**homecoming** |  |  |  | Yes | Yes |  |  |  |  |  |
| **Availability of**<br>**ongoing** | Yes |  | Yes |  | Yes | Yes | Yes | Yes |  | Yes |
| **family**<br>**support** |  |  |  |  |  |  |  |  |  |  |
| **Twins' age in**<br>**months at the**<br>**time of the**<br>**survey** | 5 | 10 | 12 | 12 | 5 | 12 | 10 | 12 | 3 | 15 |

V: vaginal delivery, CS:Caesarean section
NICU: neonatal intensive care unit
M: male, F: female

### Perceptions of twin fatherhood

The analysis using the KJ Method generated 388 labels from the interview transcripts. These were refined through 6 stages of selection (“multi-stage pick-up”) to 43 core labels, which were then integrated into 10 conceptual groups, or “thematic islands” (Supplementary Material) . Each island was assigned a symbolic theme, and their interrelationships were visualized in a structural diagram (Fig 5). The category titles (hyosatsu) generated at each stage of integration and the original labels contained within each “island” are presented in S1 Table. In the first stage of integration, any original labels that could not be integrated with others were treated as “lone wolves” and carried forward to the next stage of integration. Accordingly, Supplementary Material lists these lone labels alongside first-stage hyosatsu to clearly illustrate the correspondence. To interpret the results, a storyline was constructed based on the 10 islands identified through the KJ method, each of which was assigned a symbolic theme (referred to as a “symbol”) to represent its essence. In the main text, these symbols are presented in quotation marks ({ }).

**Fig 5.**
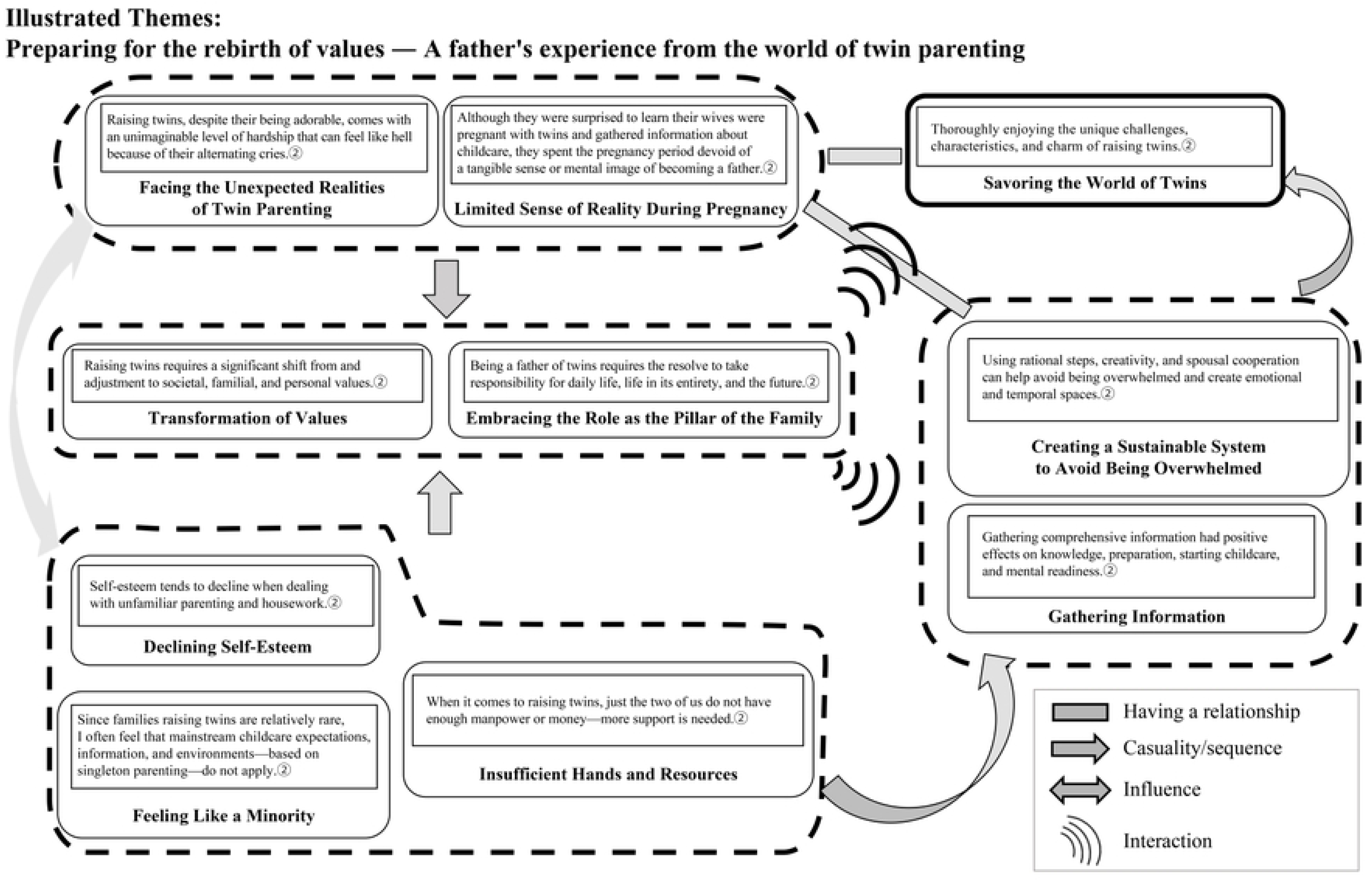
Structured Illustration of Fathers’ Perceptions of Child Care.

The following storyline depicts the psychological and emotional changes, as well as the reconstruction of paternal role awareness by fathers of twins from the pregnancy period through the child-rearing phase.

After learning of their partners’ twin pregnancies, the fathers {had only a limited sense of reality during pregnancy} regarding becoming fathers of twins. It was only after the birth of their children that they were faced with the {unexpected realities of twin parenting}. In the midst of the daily struggles and confusion of child-rearing, they { experienced a decline in self-esteem}. The physical burden of twin parenting, including the perception that {there are not enough hands or money,} was compounded by the social reality of the rareness of families raising twins, leading many to feel a sense of {minority status} within their communities. These difficulties forced the fathers to reconsider and reconstruct their social roles, positions within the family, and personal values, ultimately experiencing a {transformation of values.} In this process, a {sense of resolve as the pillar of the family} was internalized, and a proactive redefinition of their paternal roles was observed. To overcome the challenges of raising twins, the fathers found that {gathering comprehensive information} right from the pregnancy period, preparing for the confusion after birth, and coordinating rationally with their spouses were essential. They also discovered that {creating a system that prevents them from being overwhelmed by parenting can generate a sense of control.} Through repeated trial and error, the fathers eventually began to focus not only on the hardships unique to raising twins but also on the enjoyment and joy it brings. Ultimately, they arrived at a multifaceted and positive reinterpretation, expressed as {savoring the world of twins.}

From this storyline, it became clear that fathers of twins—who initially lacked a concrete understanding of their role during the pregnancy period—underwent a dynamic and developmental process of adaptation. By confronting the realities of childbirth and parenting, they experienced psychological and emotional struggles, gradually reconstructing their awareness of what it means to be a father.

The 10 symbolic themes derived from the data are outlined below. In the description, the original label is indicated by “ ”, the first stage of the integration is indicated by <>, and the second stage of name plates is indicated by 《 》.

### Limited sense of reality during pregnancy

Fathers reported that《although they were surprised to learn their wives were pregnant with twins and gathered information about childcare, they spent the pregnancy period devoid of a tangible sense or mental image of becoming a father. 》 One stated, “When I heard it was twins, instead of going blank, I was like, ‘Wow!’—it was just surprising.” “It was only after holding their child post-birth that they felt a strong realization of fatherhood. They recognized that <even if they obtained information during pregnancy, it was difficult to form a concrete image or sense of becoming a father of twins until after the birth.>

### Facing the unexpected realities of twin parenting

During pregnancy, fathers did not have a realistic sense of becoming fathers of twins.

However, after their children were discharged and parenting began, they were abruptly confronted with the reality. Fathers described, <Even though my children are cute, the alternating crying felt like hell> and <RAISING twins is harder than I could have ever imagined>. These narratives reflect how overwhelmed and beyond expectation the experience was.

### Declining self-esteem

As exhaustion from the unexpected intensity of twin parenting accumulated, fathers found that the parental role of “child rearing” often failed to produce visible outcomes, leading to a decline in self-esteem and vulnerability to mental fatigue. Even if they had believed they could manage before the birth, they came to recognize, <I’m someone whose self-esteem tends to decline when dealing with unfamiliar parenting and housework.> Statements such as, “I want my wife to occasionally praise my childcare efforts,” and “I felt ashamed and depressed when I couldn’t manage the housework well,” reflect their inner struggle.

### Insufficient hands and resources

Fathers acknowledged that 《when it comes to raising twins, just the two of us are not enough—more support is needed》. One reflected, ˂to do everything ourselves only leads to a dead end.≥ While taking paternity leave was expected, they also hoped for additional help, such as “support visits a few times a month, possibly subsidized.” Because the financial burden of raising twins falls heavily on the household, fathers expressed, “Families with twins need more financial support than do those with singletons.” Additionally, even when support systems exist, they are not always accessible in practice, as revealed by statements such as “I wanted postpartum care but couldn’t afford it,” and “You can’t schedule sudden needs—what we need is immediate support.”

### Feeling like a minority

Since families raising twins are relatively rare, fathers often 《felt that mainstream childcare expectations, information, and environments—based on singleton parenting—did not apply.》 For instance, they noted, “The booklet from the city only had information for one child,” and “Most public facilities are built with only one child in mind.” When trying to explain the hardships of raising twins, they found that “the specific difficulty of having twins isn’t understood by those raising just one child,” creating a sense of being misunderstood.

### Transformation of values

Fathers recognized that 《raising twins required a significant shift and adjustment in societal, familial, and personal values.》 Looking to the future, they stated, “To build a relationship with my children, I need to spend time with them,” and “How much time I can give depends on workplace understanding.” They also expressed, “During the twin parenting period, it’s necessary to rethink how one lives in society.”

### Embracing the role as the pillar of the family

While it was difficult to imagine becoming a father of twins during pregnancy, through parenting and household responsibilities, fathers gradually realized that 《being a father of twins requires the resolve to take responsibility for daily life, the entirely of life, and the future.》 One described <THE fear of bearing the burden alone when told by doctors about perinatal complications> and another recalled the moment of discharge: <When the hospital handed over the children and said, ‘Here you go,’ it felt like they were passing the baton of life.> These experiences helped fathers develop a profound sense of responsibility as the central figure supporting the family.

### Gathering comprehensive information

Although it was difficult to envision or internalize the role of a twin father during pregnancy, fathers understood that “gathering comprehensive information had positive effects on knowledge, preparation, starting childcare, and mental readiness.” They stated, “Without basic knowledge about twin pregnancy and childbirth, I couldn’t keep up with the conversation with the doctor or my wife when complications occurred” and “Understanding changes in my wife’s mental health before and after childbirth helped her a lot” They felt that <summarizing what fathers can do during pregnancy through information gathering and rehearsals made starting childcare smoother.> Some participated in twin-specific parenting classes, practiced household chores and caregiving using dolls, or listed what they could do from pregnancy onward. Additionally, fathers suggested that “it would be helpful to include voices from experienced twin fathers in materials” and expressed a desire for “opportunities to talk with other twin fathers.” These voices clearly indicate a need for peer support from those with similar experiences.

### Creating a sustainable system to avoid being overwhelmed

Despite the severity of the parenting experience, fathers discovered that <by using rational steps, creativity, and spousal cooperation, it is possible to avoid being overwhelmed and to create emotional and temporal spaces>. They believed that “coordination between spouses is key,” and emphasized the need to share information efficiently even under time constraints. They “shared strategies such as synchronizing feeding cycles, so both babies cry together, as well as rotating breastfeeding, formula, and pumping.” One father stated, “Treating parenting like a set of tasks made it easier.” They also found that by maintaining an active awareness of how to manage time, they could <secure personal time through deliberate control.>

### Savoring the world of twins

Fathers acknowledged that they were《thoroughly enjoying the unique challenges, characteristics, and charm of raising twins.》 They became aware that “even when interacting equally, the children’s responses differ,” and “it’s difficult to understand why each baby cries—each has different needs.” One father noted, “Although I felt differently about each child’s personality, I didn’t worry about it,” while another said, “There’s a communication between twins that parents can’t fully understand.” As one father summed up, “Raising twins is twice as hard, but the joy and happiness from watching them grow is also double,” reflecting the depth of meaning they found in this unique parenting experience.

## Discussion

This qualitative study explored how fathers of twins acquire paternal roles and redefine the burdens of parenting under the unique conditions of raising twins. Based on Thornton and Nardi’s role acquisition model [17], it focused on the early parenting period—from pregnancy to approximately one year postpartum—and applied the KJ method to structurally analyze fathers’ subjective narratives. The three key contributions are as follows:

First, this study visualized the process of cognitive transformation among fathers of twins— particularly how they acquired paternal identity under unique stressors—by structurally organizing their experiences from pregnancy onward. Second, it revealed how intrinsic motivation developed through fathers’ experiences of psychological exhaustion—including decreased self-efficacy—and how they restructured their parenting environments through spousal cooperation and practical innovations. Third, it identified the psychosocial stressors specific to twin-parenting families, such as a sense of minority status, social system inadequacies, and limited regional resources—all of which deeply influence how fathers redefined their paternal roles.

Previous research has mainly focused on fathers of singletons, documenting their paternal role development and involvement in childcare [37]. However, qualitative studies that analyze the cognitive transformation process of fathers of twins—especially from the prenatal period through the early stages of child rearing—are notably scarce [5,6].

Our findings revealed that fathers, upon the birth of their twins, were forced to reassess their prior values and roles. Faced with overwhelming realities beyond expectations, many experienced decreased self-esteem and mental distress. Nevertheless, by collaborating with their spouses and restructuring their approach to parenting, they gradually developed intrinsic motivation and began reframing their experiences positively—eventually expressing joy in “immersing themselves in the world of twins.” This finding also aligns with previous findings regarding turning points in paternal burden [16].

Another important insight is the emergence of “paternal resolve” as described by the fathers.

They did not merely perceive themselves as supporters of the family but as core figures who “take responsibility for the entirety of life.” This represents a redefinition of fatherhood and a departure from conventional gender norms, signaling the emergence of a new fatherhood identity [38]. Fathers’ proactive efforts during pregnancy, including gathering medical information and preparing for perinatal risks, also reflect a broader trend of men’s increased involvement in care and parenting [11].

Institutional insights also emerged. Fathers often expressed a sense of social marginalization (“minority feeling”) and frustration with child-rearing policies largely modeled on singleton families. Difficulties in accessing childcare leave, insufficient public support systems, and gaps in service design contributed to stress and hindered the development of fatherhood [39,40].

From a life-course perspective [41], many fathers were in midlife—a stage associated with growing professional responsibilities and personal development. The pressure to fulfill parenting duties alongside career demands, concerns about workplace evaluation, and lack of understanding from superiors and colleagues were all identified as external factors influencing their psychological adjustment [42].

These findings highlight the importance of establishing support mechanisms for fathers of twins from the early prenatal stage. These should include disseminating information tailored to twin pregnancies, promoting peer-support programs, and implementing workplace reforms allow flexible paternal leave. Furthermore, policy efforts should explicitly include twin-parenting families as a distinct group requiring targeted support—not as exceptional cases but as part of standard childcare planning.

## Conclusions

This study described the process by which fathers of twins, despite experiencing diminished self-efficacy, as well as physical and emotional exhaustion in the early stages of parenting, reconstructed their childcare environment through spousal collaboration and creative strategies, eventually discovering a unique “meaning” in raising twins, transitioning from the “anticipatory” to the “personal” stage in Thornton & Nardi’s role acquisition model, with the unique conditions of twin parenting catalyzing fathers’ internal growth and active acquisition of paternal roles. In particular, the realization of their commitment as the “central figure” in the family and their proactive engagement in childcare beyond the bounds of traditional social and cultural norms provide important insights into the redefinition of fatherhood.

The findings underscore the need for early interventions to support fathers and contribute to the development of comprehensive support systems that consider the complexities of parenting twins. In the future, phased support programs for fathers beginning in the prenatal period should be developed to promote broader social understanding involving families, communities, and workplaces, thereby fostering environments that enable sustainable work–life balance for fathers.

### Limitations

This study has several limitations. First, the participants were limited to fathers raising healthy twins. Families of children with developmental disorders, medical conditions, or sequelae from premature birth were not included. Therefore, because the findings cannot be generalized to those contexts, future research should include a more diverse range of participants. Second, the participants experienced childbirth and parenting during the COVID-19 pandemic, under conditions that included restricted hospital visitation and limited access to external support, conditions that may have posed unique challenges to the process of acquiring paternal roles and receiving parenting support. Third, the participants were drawn from specific geographic regions (the Tokyo and Kansai metropolitan areas) and had relatively homogeneous socioeconomic backgrounds in terms of occupation and education. Thus, the findings may not fully reflect national trends or cultural variations. Furthermore, owing to the online nature of data collection, nonverbal expressions and embodied interactions could not be fully captured.

In light of these limitations, future studies should involve fathers with more diverse attributes and incorporate comprehensive and comparative perspectives that consider different parenting environments and cultural contexts.

## Data Availability

The data cannot be shared publicly because they contain personally identifiable information of study participants. Data are available from the Research Ethics Committee of Kansai Medical University for researchers who meet the criteria for access to confidential data.

## Acknowledgments

We extend our sincere gratitude to all the fathers who participated in this study. We also express our deep appreciation to the institutions and individuals who supported the study by facilitating participant recruitment and understanding the purpose of the research. This study is based on a doctoral dissertation submitted to the Graduate School of Nursing, Kansai Medical University, and has been revised and expanded for publication.

